# LiteMIL: A Computationally Efficient Transformer-Based MIL for Cancer Subtyping on Whole Slide Images

**DOI:** 10.1101/2025.05.11.25327389

**Authors:** Haitham Kussaibi

## Abstract

**Purpose:** Accurate cancer subtyping is crucial for effective treatment; however, it presents challenges due to overlapping morphology and variability among pathologists. Although deep learning (DL) methods have shown potential, their application to gigapixel whole slide images (WSIs) is often hindered by high computational demands and the need for efficient, context-aware feature aggregation. This study introduces LiteMIL, a computationally efficient transformer-based multiple instance learning (MIL) network combined with Phikon, a pathology-tuned self-supervised feature extractor, for robust and scalable cancer subtyping on WSIs.

**Methods:** Initially, patches were extracted from TCGA-THYM dataset (242 WSIs, six subtypes) and subsequently fed in real-time to Phikon for feature extraction. To train MILs, features were arranged into uniform bags using a chunking strategy that maintains tissue context while increasing training data. LiteMIL utilizes a learnable query vector within an optimized multi-head attention module for effective feature aggregation. The model’s performance was evaluated against established MIL methods on the Thymic Dataset and three additional TCGA datasets (breast, lung, and kidney cancer).

**Results:** LiteMIL achieved 0.89 ± 0.01 F1 score and 0.99 AUC on Thymic dataset, outperforming other MILs. LiteMIL demonstrated strong generalizability across the external datasets, scoring the best on breast and kidney cancer datasets. Compared to TransMIL, LiteMIL significantly reduces training time and GPU memory usage. Ablation studies confirmed the critical role of the learnable query and layer normalization in enhancing performance and stability.

**Conclusion:** LiteMIL offers a resource-efficient, robust solution. Its streamlined architecture, combined with the compact Phikon features, makes it suitable for integrating into routine histopathological workflows, particularly in resource-limited settings.

## 1. INTRODUCTION

Accurate subtyping of cancers is a cornerstone of precision medicine, directly influencing therapeutic decisions and prognostic assessments. However, the histopathological evaluation remains fraught with challenges, including substantial morphological overlap between tumor subtypes and high inter-observer variability among pathologists. Convolutional neural networks (CNNs) have demonstrated remarkable performance in various medical imaging tasks, including tumor detection, grading, and subtyping. In histopathology, CNN-based models have shown promise in automating complex diagnostic processes and reducing observer bias. The digital revolution in pathology has enabled the routine acquisition and storage of gigapixel of whole slide images (WSIs), opening new avenues for computational analysis and the development of artificial intelligence (AI)-driven diagnostic tools (1-6). Despite these advances, the translation of deep learning (DL) models into clinical practice is hindered by WSI analysis’s unique technical and practical demands. The direct application of CNNs to WSIs is computationally prohibitive due to the sheer size of these images, which can exceed 100,000 × 100,000 pixels. This necessitates patch-based approaches, where WSIs are divided into smaller, manageable tiles for analysis. While patch-based methods alleviate computational burdens, they often sacrifice the broader tissue context essential for nuanced cancer subtyping, potentially limiting diagnostic accuracy (7). To address these limitations, Multiple Instance Learning (MIL) has emerged as a robust and flexible framework for WSI analysis (7-9). In MIL, each WSI is treated as a bag of instances (patches), and only slide-level labels are required for supervision, circumventing the need for exhaustive manual annotation (10-13). The seminal attention-based MIL (ABMIL) framework by Ilse et al. (14) introduced attention mechanisms to weight instances according to their diagnostic relevance, significantly improving the identification of critical regions within WSIs. Subsequent enhancements have refined instance selection through various mechanisms (14-17).

Multi-head attention-based MIL (MAD-MIL) has further refined instance selection and improved classification performance by leveraging parallel attention heads (18). However, attention-based methods do not capture the morphology context in WSIs. Transformer-based MIL models, such as TransMIL, have leveraged self-attention mechanisms combined with positional encoding to capture complex inter-instance dependencies, achieving state-of-the-art results in WSI classification (19, 20). Despite their improved performance, transformer-based MILs often suffer from high computational overhead, particularly when processing bags containing thousands of instances, as is typical in WSI analysis (19, 20). The quadratic complexity of self-attention mechanisms concerning the number of instances can lead to excessive memory consumption and long training times, posing significant barriers to clinical deployment, especially in resource-constrained settings. This prompts the development of more efficient alternatives that streamline transformer-based methods while maintaining robust performance. In this context, we propose a computationally efficient MIL network (LiteMIL), LiteMIL, a Streamlined Transformer-based MIL Framework, is a novel MIL architecture that employs a dedicated learnable query vector within a built-in optimized multi-head attention module for efficient global feature aggregation. By replacing full self-attention with a query-driven aggregation mechanism, LiteMIL significantly reduces memory usage and training time compared to traditional transformer-based MILs, like TransMIL, without sacrificing diagnostic performance (19, 20). Ablation studies demonstrate that the learnable query and layer normalization are critical for LiteMIL robustness and accuracy. We comprehensively evaluated LiteMIL on the TCGA-THYMIC dataset, which poses a challenging multi-class subtyping task with significant morphological overlap.

We also benchmark LiteMIL’s generalizability across three external cancer cohorts, adhering to best practices for reproducible computational pathology research. Our results show that LiteMIL achieves state-of-the-art accuracy while significantly reducing computational requirements compared to transformer-based MIL, making it well-suited for integration into clinical workflows, especially in resource-limited settings.

Feature extraction is another critical determinant of DL model performance in computational pathology. Traditionally, CNNs pre-trained on natural image datasets such as ImageNet have been widely adopted for feature extraction in histopathology (21). However, these models may not fully capture the subtle morphological variations unique to pathology images, potentially limiting their discriminative power (22). Recent progress in self-supervised learning (SSL) has enabled the development of domain-specific feature extractors, tailored to the visual characteristics of histopathological images, generating compact, highly discriminative representations.

SSL-based models have demonstrated improved classification accuracy and computational efficiency (23-26). Herein, we used Phikon, a ViT-based self-supervised learning model optimized for histology images, which generates compact 768-dimensional feature vectors from WSI patches (23). To efficiently manage the vast number of feature vectors per WSI, we employ a chunking strategy that partitions each WSI into multiple, uniformly sized bags (e.g., 1,000 vectors per bag). This approach augments the training set while preserving local tissue context. Unlike random subsampling, this method ensures comprehensive coverage of WSI while simplifying batch processing and reducing the need for complex padding or masking operations.

In this study, combining LiteMIL, Phikon, and optimized feature bagging enhanced diagnostic performance and resource utilization. By addressing the dual challenges of diagnostic precision and computational efficiency, our work contributes to ongoing efforts to translate artificial intelligence from research into routine clinical practice in digital pathology.

## 2. METHODS

This section details dataset preparation (including patch extraction, feature extraction, and organization), LiteMIL architecture design, training protocols, and evaluation strategies. The whole method was evaluated on TCGA-THYM WSIs, while three ready-to-use feature datasets, prepared by MAD-MIL study (18), were used to assess LiteMIL’s performance and generalizability.

### 2.1 THYMIC DATASET PREPARATION

We utilized the publicly available TCGA-THYM dataset, comprising 124 cases (61 females, 63 males; age range: 61–101 years), totaling 242 WSIs after excluding slides of poor quality or ambiguous diagnosis. Tumors were categorized into six subtypes according to the 2021 WHO classification: A, AB, B1, B2, B3, and carcinoma (27).

#### 2.1.1 Data Splitting

To ensure balanced representation, WSIs were partitioned into training and test sets using an 80:20 stratified split, implemented via Scikit-learn’s StratifiedKFold (28). The test set was reserved exclusively for final evaluation to prevent data leakage.

#### 2.1.2 Patch Extraction

Given the gigapixel scale of WSIs, we employed a customized method derived from the Yottixel search engine for patch extraction (29). It creates masks to isolate patches exclusively from tissue regions with up to a 30% background-to-tissue ratio, discarding patches that exceed the threshold. The extracted patches are then fed in real time to a feature extraction network without intermediate storage.

#### 2.1.3 Feature Extraction

Two feature extraction models, an SSL model and a traditional CNN, were compared:

##### Phikon

A ViT-based SSL model optimized for histology images (23), generating 768-dimensional feature vectors. Phikon includes its own preprocessing pipeline.

##### ResNet50

A conventional CNN Pre-trained on ImageNet (21), and fine-tuned on the UBC-OCEAN dataset (30), producing 2048-dimensional feature vectors.

All extracted features were normalized prior to downstream processing.

#### 2.1.4 Bag Formation via Chunking

We employed a chunking strategy to divide each WSI’s feature set into evenly sized bags (usually 1,000 instances per bag). For instance, a WSI containing 8000 feature vectors will be divided into eight bags. This method augments the training set, retains local tissue context, simplifies batch processing, and lowers the computational load of analyzing the entire WSI as one bag.

### 2.2 LITEMIL ARCHITECTURE DESIGN (SEE Fig. 1)

The LiteMIL framework consists of four main modules:

1. **Feature Projection Module**: Each patch-level feature is projected via a linear layer, followed by Layer Normalization, ReLU activation, and dropout, yielding a latent representation.
2. **Learnable Query Vector:** A single learnable query, expanded to batch size, serves as the sole query for the self-attention-based aggregation.
3. **Multi-Head Attention Aggregation**: PyTorch’s nn.MultiheadAttention (17) is used, with the learnable query attending to all projected features, producing a bag-level representation. This design reduces the computational complexity compared to full self-attention (19).
4. **Classifier Module**: The aggregated bag-level features are passed through a multi-layer perceptron (MLP) with GELU activation and dropout, outputting logits for subtype classification.

#### 2.2.1 Ablation Studies

To assess the contribution of individual architectural components, we conducted ablation experiments varying dropout, Layer Normalization, the learnable query vector, the number of attention heads, and other parameters. Performance was evaluated in terms of F1-score and training stability.

### 2.3 EXTERNAL VALIDATION DATASETS

To evaluate generalizability, we used three ready-to-use feature datasets prepared by MAD-MIL project (18):

- **TUPAC16:** 821 WSIs from TCGA breast cancer, labeled as low-grade or high-grade.
- **TCGA Lung Cancer:** 1,046 WSIs labeled as adenocarcinoma or squamous cell carcinoma.
- **TCGA Kidney Cancer:** 918 WSIs labeled as Clear Cell, Papillary, or Chromophobe Renal Cell Carcinoma.

### 2.4 MODEL TRAINING & EVALUATION

#### 2.4.1 Experimental Setup

Training was performed on a workstation with an NVIDIA GPU (12 GB, 8,000 CUDA cores), 32 GB RAM, and an Intel Core i7 13th Gen processor. All experiments used PyTorch (v1.12.1). Key hyperparameters are summarized in Table 1.

**Table 1:**
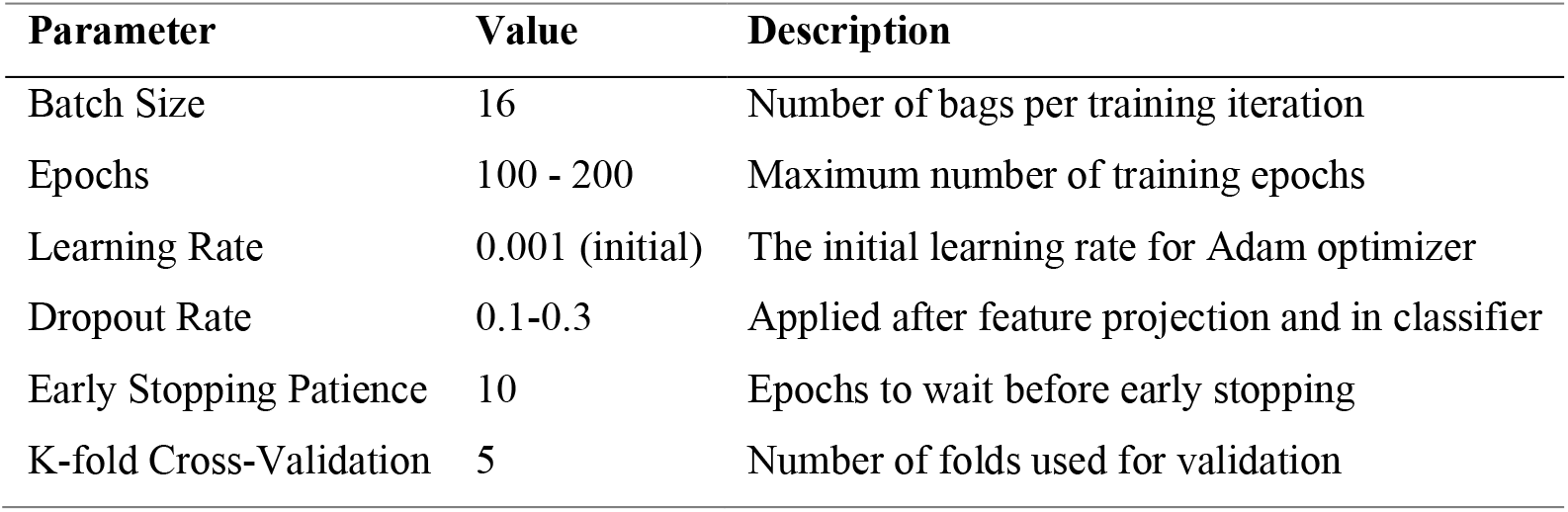
Optimized key hyperparameters.

#### 2.4.2 Training Protocol

- **Optimizer**: Adam, with a learning rate scheduler (patience = 3).
- **Early Stopping**: Monitored validation loss to prevent overfitting.
- **Loss Function**: Customized class-weighted cross-entropy to address class imbalance.
- **Cross-Validation**: 5-fold, with fixed random seeds for reproducibility.

#### 2.4.3 Evaluation Metrics

- **F1-Score**: Computed per subtype and as a weighted average.
- **AUC-ROC**: One-vs-rest (OvR) for multiclass tasks.
- **Confusion Matrix**: To visualize misclassification patterns.
- **Computational Efficiency**: GPU memory usage and training time were recorded.

All results are reported as mean ± standard deviation across multiple runs/folds.

## 3 RESULTS

### 3.1 THYMIC Dataset Summary and Patch Extraction

From the 242 WSIs in the TCGA-THYM dataset, the Yottixel patch extraction method yielded a total of 428,000 tissue patches (each 250×250 pixels). These patches were processed in real-time by both Phikon and ResNet50 feature extractors, resulting in 428 feature bags per extractor (see Table 2 for subtype distribution).

**Table 2:** Distribution of WSIs, patches, and bags across thymic tumor subtypes.

| Subtype | N. of WSIs | N. of Patches | N. of Bags<br>(1k-size) |
| --- | --- | --- | --- |
| A | 34 | 56k | 56 |
| AB | 68 | 120k | 120 |
| B1 | 39 | 80k | 80 |
| B2 | 61 | 102k | 102 |
| B3 | 22 | 40k | 40 |
| CA | 18 | 30k | 30 |
| Total | 242 | 428k | 428 |

#### 3.1.1 Impact of Bag Size on Performance and Efficiency

Experiments with varying bag sizes (200, 500, and 1,000 patches per bag) using the Phikon-LiteMIL network demonstrated that 1,000-patch bags provided the optimal balance between context preservation and data augmentation. Smaller bag sizes increased the number of training samples but led to reduced performance, likely due to insufficient contextual information. The bag configuration also minimized GPU memory usage and training time (Table 3).

**Table 3:**
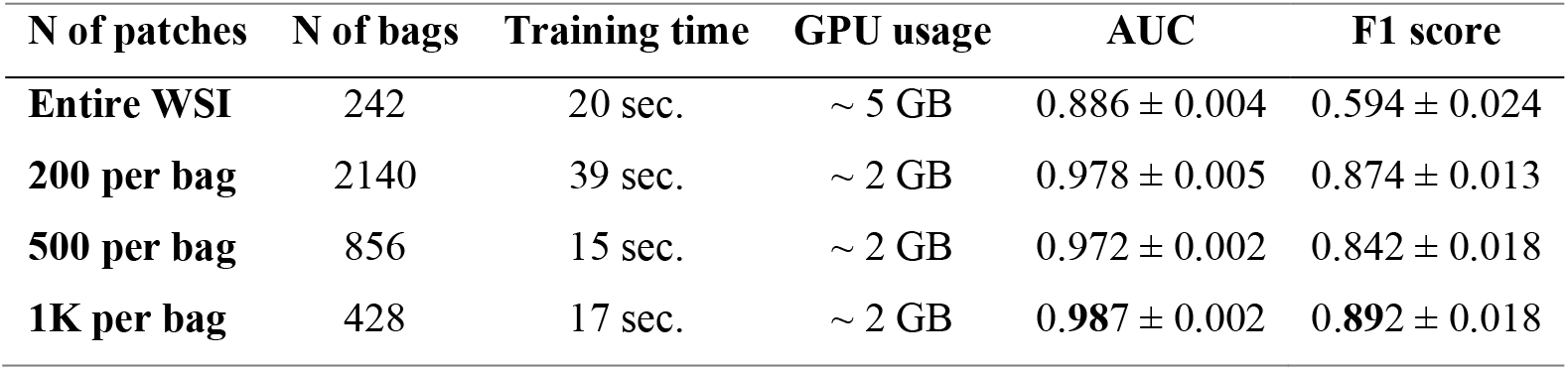
Impact of bag size on performance and computational efficiency.

### 3.2 ABLATION STUDY OF LITEMIL ARCHITECTURE

Ablation experiments assessed the contribution of key architectural components in LiteMIL (tested on the Phikon-extracted thymic dataset). Incorporating a learnable query vector improved model robustness and reduced performance variance. Layer Normalization stabilized feature scaling and further enhanced performance, while mild dropout contributed positively to generalization. The use of ReLU activation in the feature projection module and GELU in the MLP classifier also improved results (Table 4).

**Table 4:** Ablation study results for LiteMIL components.

| Model Variation | F1 score (± STD) |
| --- | --- |
| LiteMIL Model | **$0.89 \pm 0.01$** |
| Without Dropout | $0.87 \pm 0.02$ |
| Without Layer Normalization | $0.86 \pm 0.03$ |
| Without Attention Mechanism | $0.87 \pm 0.03$ |
| Linear Classifier head | $0.87 \pm 0.02$ |
| ReLU → GELU (in MLP) | $0.86 \pm 0.02$ |
| Without Learnable Query | $0.87 \pm 0.04$ |
| Attention Heads (2 instead of 4) | $0.87 \pm 0.02$ |

### 3.3 PERFORMANCE COMPARISON: PHIKON VS. RESNET50 FEATURES

LiteMIL achieved an F1 score of 0.892 ± 0.018 and an AUC of 0.987 ± 0.002 on the Phikon-extracted thymic dataset, outperforming its performance on the ResNet50-extracted dataset (F1: 0.703 ± 0.017, AUC: 0.915 ± 0.007; see Table 5). Training and validation curves (Fig. 2) showed that LiteMIL-ResNet50 struggled to converge and exhibited signs of overfitting, while LiteMIL-Phikon maintained stable learning dynamics. **Confusion matrices** (Fig. 3) revealed that the Phikon-LiteMIL model achieved perfect recall (1.00) for carcinoma (CA) and B1 subtypes, and high precision (>90%) for A and B3 subtypes. Misclassifications were most frequent between AB, B1, and B2 subtypes, reflecting known morphological overlaps. In contrast, ResNet50-LiteMIL showed lower performance across most subtypes, particularly for CA and B2.

**Table 5:**
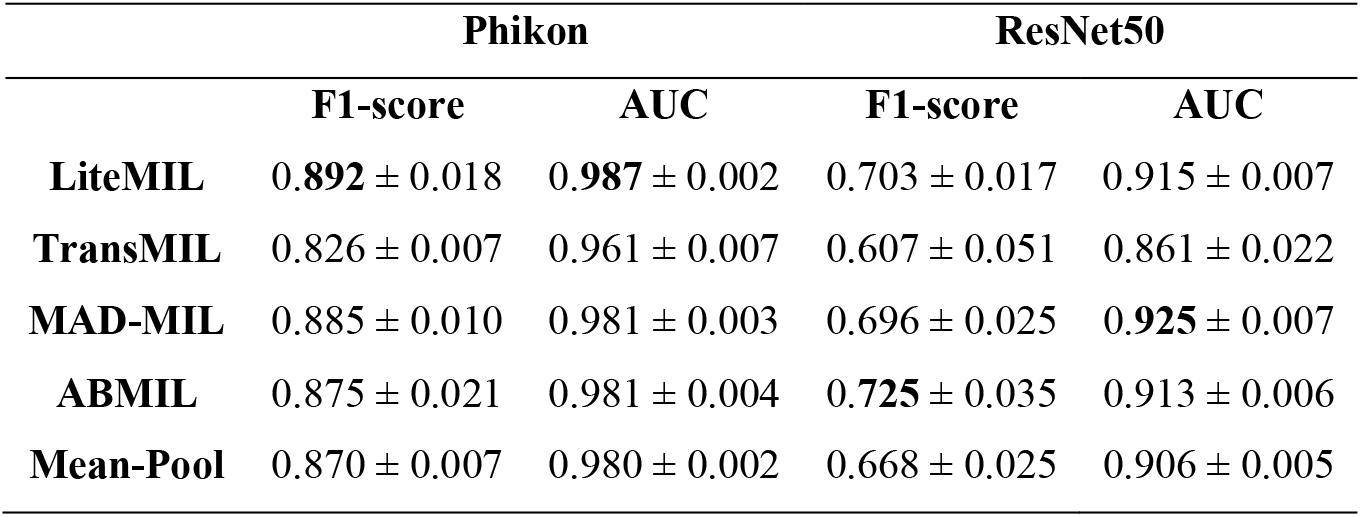
F1-score and AUC of extractor-classifier combinations on the thymic dataset.

|  | Phikon |  | ResNet50 |  |
| --- | --- | --- | --- | --- |
|  | F1-score | AUC | F1-score | AUC |
| **LiteMIL** | **0.892** ± 0.018 | **0.987** ± 0.002 | 0.703 ± 0.017 | 0.915 ± 0.007 |
| **TransMIL** | 0.826 ± 0.007 | 0.961 ± 0.007 | 0.607 ± 0.051 | 0.861 ± 0.022 |
| **MAD-MIL** | 0.885 ± 0.010 | 0.981 ± 0.003 | 0.696 ± 0.025 | **0.925** ± 0.007 |
| **ABMIL** | 0.875 ± 0.021 | 0.981 ± 0.004 | **0.725** ± 0.035 | 0.913 ± 0.006 |
| **Mean-Pool** | 0.870 ± 0.007 | 0.980 ± 0.002 | 0.668 ± 0.025 | 0.906 ± 0.005 |

**Fig. 1:**
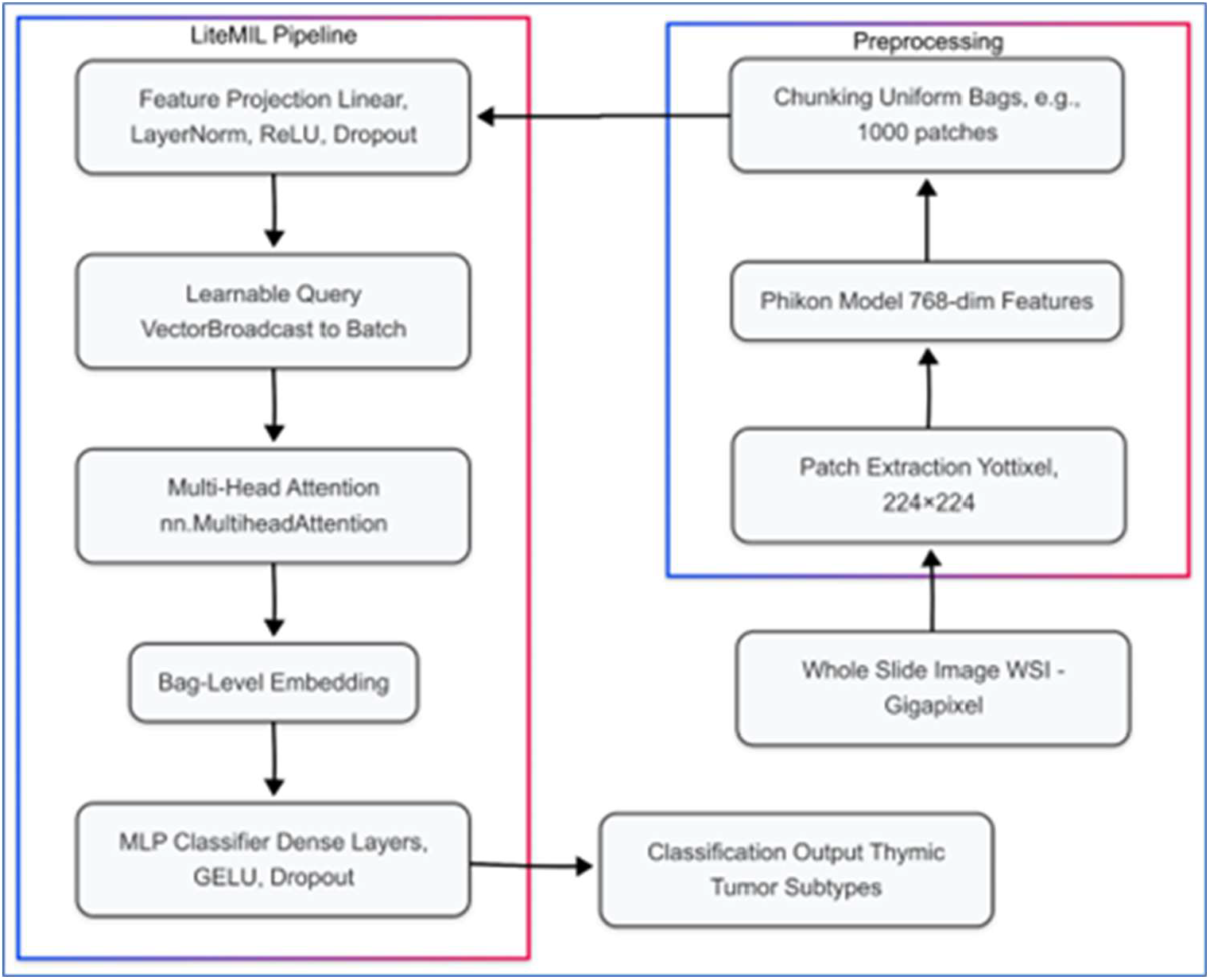
The workflow of cancer subtyping from WSI to Classification output.

**Fig. 2:**
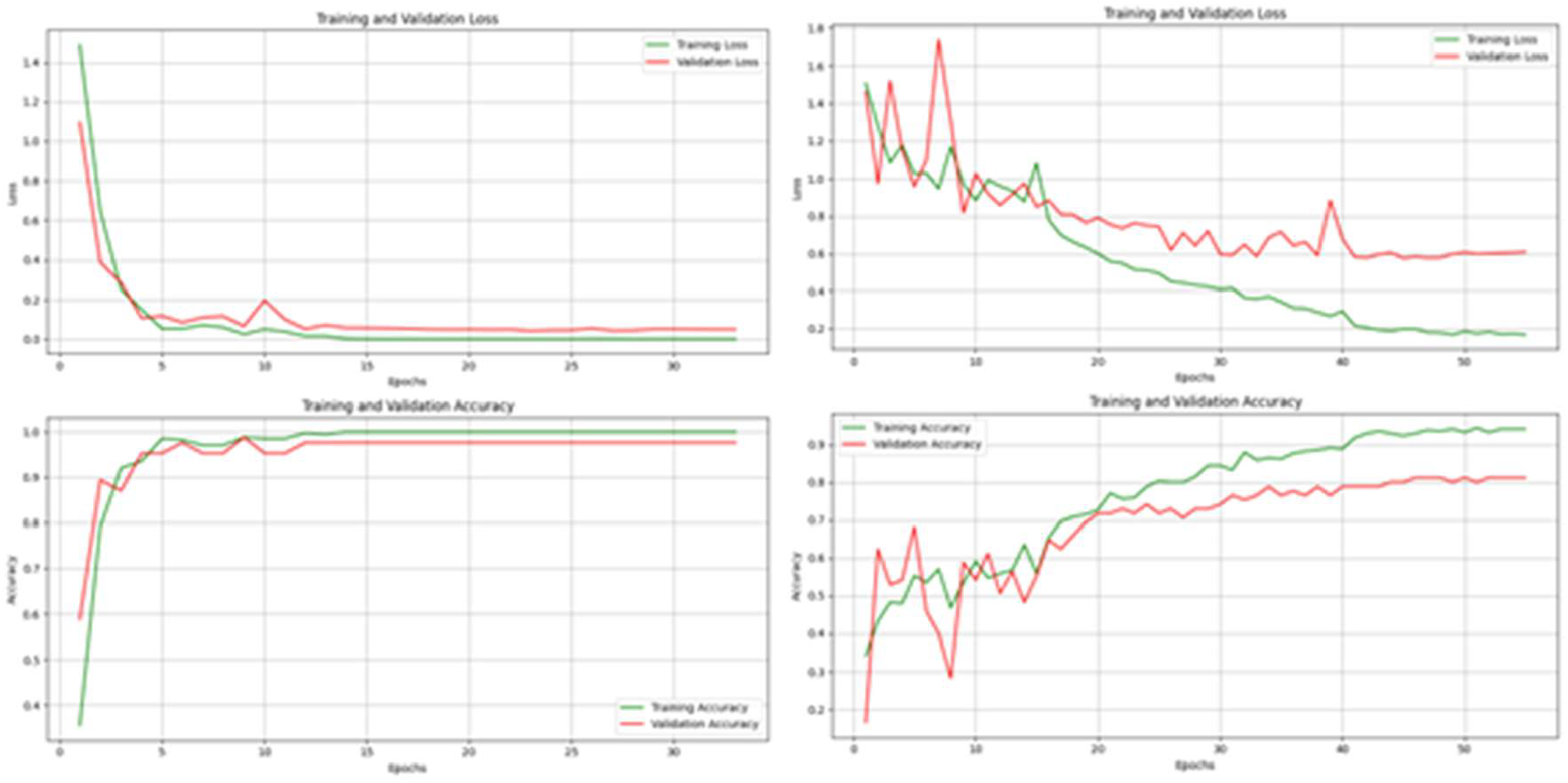
Training and validation performance curves for LiteMIL-Phikon (A) and LiteMIL-ResNet50 (B).

**Fig. 3:**
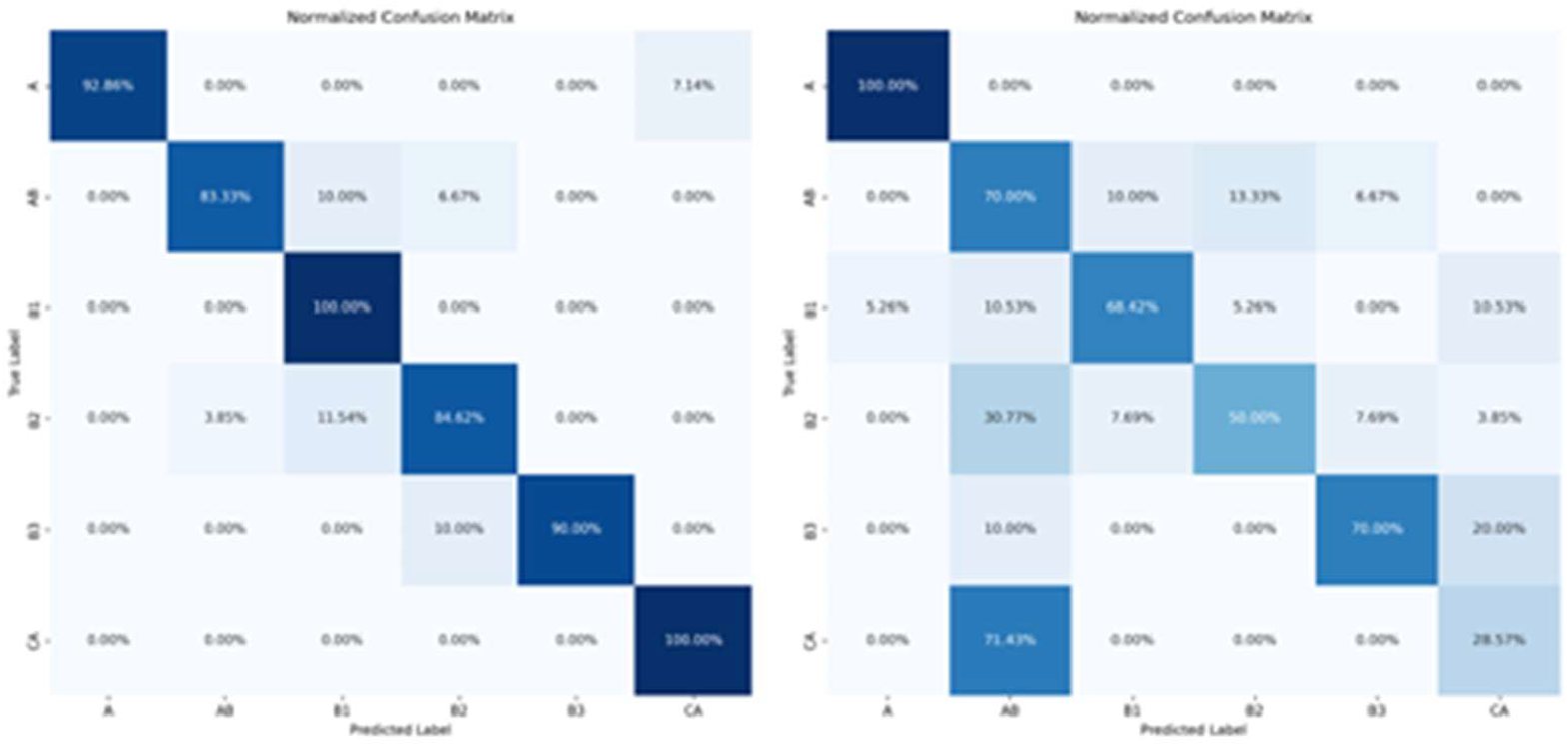
Confusion matrix for the Phikon-LiteMIL (A) and ResNet50-LiteMIL (B) models.

### 3.4 COMPARISON WITH ESTABLISHED MIL METHODS

#### 3.4.1 Performance on Thymic Dataset

On the Phikon-extracted thymic dataset, LiteMIL outperformed benchmarked MIL methods, and mean-pool classifier in both F1 score and AUC (Table 5). On the ResNet50-extracted dataset, LiteMIL’s performance was comparable, better, or slightly lower than other methods.

#### 3.4.2 Performance on External Datasets (Generalizability)

LiteMIL was evaluated on three external TCGA-derived ready-to-use feature datasets (TUPAC16, Lung, and Kidney). The model achieved the highest F1 scores on the TUPAC16 (breast cancer) and kidney cancer datasets, and competitive performance on the lung cancer dataset (Table 6). These results demonstrate strong generalizability across diverse histopathology cohorts.

**Table 6:**
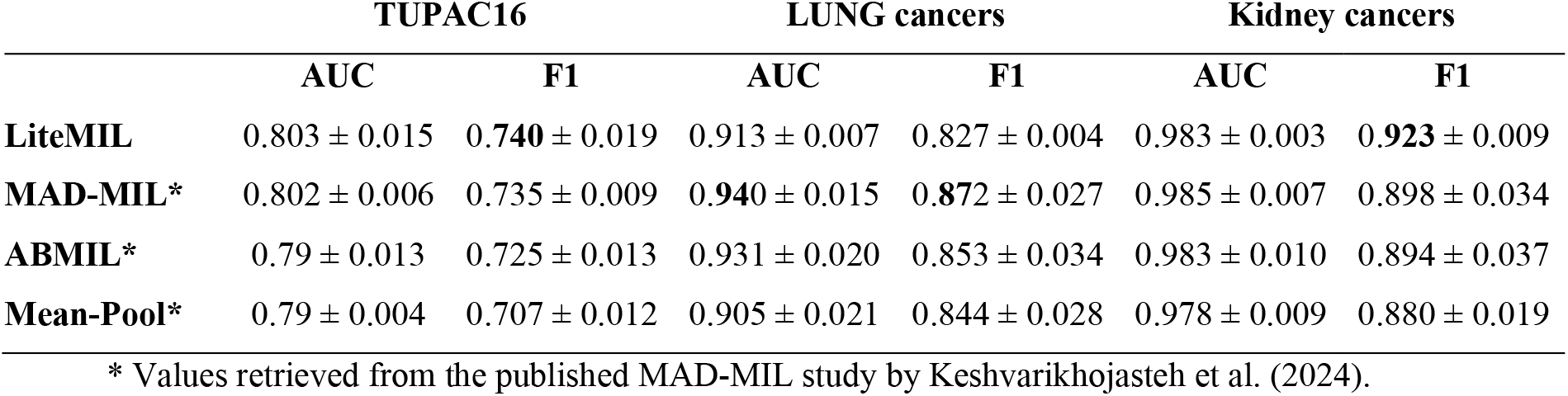
LiteMIL performance on external datasets compared to other MIL methods.

|  | TUPAC16 |  | LUNG cancers |  | Kidney cancers |  |
| --- | --- | --- | --- | --- | --- | --- |
|  | AUC | F1 | AUC | F1 | AUC | F1 |
| **LiteMIL** | 0.803 ± 0.015 | **0.740** ± 0.019 | 0.913 ± 0.007 | 0.827 ± 0.004 | 0.983 ± 0.003 | **0.923** ± 0.009 |
| **MAD-MIL\*** | 0.802 ± 0.006 | 0.735 ± 0.009 | **0.940** ± 0.015 | **0.872** ± 0.027 | 0.985 ± 0.007 | 0.898 ± 0.034 |
| **ABMIL\*** | 0.79 ± 0.013 | 0.725 ± 0.013 | 0.931 ± 0.020 | 0.853 ± 0.034 | 0.983 ± 0.010 | 0.894 ± 0.037 |
| **Mean-Pool\*** | 0.79 ± 0.004 | 0.707 ± 0.012 | 0.905 ± 0.021 | 0.844 ± 0.028 | 0.978 ± 0.009 | 0.880 ± 0.019 |

\* Values retrieved from the published MAD-MIL study by Keshvarikhojasteh et al. (2024).

#### 3.4.3 Computational Efficiency

Benchmarking against TransMIL on identical hardware and datasets, LiteMIL significantly reduced GPU memory usage and shortened training time, while maintaining or exceeding classification performance (Table 7). This efficiency was attributed to using a learnable query vector and optimized multi-head attention instead of full self-attention mechanisms.

**Table 7:** Computational efficiency comparison between LiteMIL and TransMIL.

| Model | LiteMIL |  | TransMIL |  |
| --- | --- | --- | --- | --- |
| Dataset | Thymic | TUPAC | Thymic | TUPAC |
| Training Time | 17 sec. | 9 min. | 17 min. | 1h51min |
| Peak GPU | 2 GB | 9 GB | 7 GB | 12 GB |
| AUC | 0.987 ± 0.002 | 0.807 ± 0.009 | 0.961 ± 0.007 | 0.760 ± 0.049 |
| F1 score | 0.892 ± 0.018 | 0.744 ± 0.015 | 0.826 ± 0.007 | 0.697 ± 0.045 |

## 4. DISCUSSION

This study presents LiteMIL, a computationally efficient transformer-based MIL framework, integrated with a pathology-tuned SSL feature extractor (Phikon), for robust cancer subtyping on WSIs. Our results demonstrate that LiteMIL achieves high diagnostic accuracy, strong generalizability across diverse cancer types, and substantial reductions in computational resource requirements compared to established transformer-based MILs. The primary finding is that LiteMIL outperforms traditional MIL methods (such as ABMIL and MAD-MIL) and the transformer-based TransMIL on the challenging Thymic feature dataset (Phikon-extracted). The F1 score of 0.89 ± 0.01 and AUC of 0.99 highlight the model’s ability to accurately distinguish between thymic tumor subtypes, including those with significant morphological overlap. Notably, LiteMIL achieved perfect recall for CA and B1 subtypes and high precision for A and B3, while most misclassifications occurred in AB and B2, mirroring the diagnostic challenges faced by human pathologists. This suggests that the model is accurate and sensitive to the nuanced patterns that define clinically relevant subtypes.

Generalizability is a critical requirement for clinical AI tools. LiteMIL demonstrated strong performance on three external TCGA datasets (TUPAC16, lung, and kidney cancers), achieving the highest F1 scores on the TUPAC16 and kidney cancer datasets and competitive results on lung cancer, despite variations in tissue type, staining, and cohort characteristics. These results support its potential for broader clinical adoption.

### 4.1 LITEMIL’S UNIQUE ARCHITECTURE VS. SIMILAR MIL METHODS

The LiteMIL framework introduces several architectural innovations that distinguish it from established MIL methods in computational pathology. Traditional MIL approaches, such as Attention-based MIL (ABMIL), rely on a self-attention mechanism where instance-level attention weights are learned through a fully connected layer, typically followed by a Tanh activation. This mechanism enables the model to assign higher importance to diagnostically relevant patches within a bag, but it aggregates information using a single attention head and a relatively shallow architecture. While effective, ABMIL may be limited in capturing complex, multi-faceted relationships among instances, especially in cases of significant morphological overlap between tumor subtypes.

MAD-MIL extends ABMIL by introducing a custom multi-head attention mechanism, where multiple sequential attention blocks, each with its own Tanh activation, are used to capture diverse aspects of the input data. The outputs of these attention heads are concatenated and passed through a linear layer for final classification. This design allows MAD-MIL to model more complex inter-instance relationships and has been shown to improve performance on challenging histopathology datasets. However, the sequential nature of its attention heads and the manual aggregation of their outputs can increase computational complexity and may not fully leverage the parallelization capabilities of modern deep learning frameworks.

In contrast, LiteMIL adopts a transformer-inspired architecture that leverages a single learnable query vector within a built-in multi-head attention module. This design enables efficient global aggregation of patch-level features by allowing the query to simultaneously attend to all instances in the bag. Unlike ABMIL, which independently computes attention weights for each instance, LiteMIL’s query-driven attention mechanism provides a more flexible and expressive means of summarizing the diagnostic content of a WSI. Multiple attention heads allow the model to capture a broader range of discriminative features, which is particularly advantageous when subtypes share overlapping morphological characteristics.

Compared to MAD-MIL, LiteMIL’s use of PyTorch’s native multi-head attention implementation streamlines the architecture, reducing the need for custom sequential blocks and manual concatenation. This not only simplifies the model but also enhances computational efficiency by fully utilizing hardware acceleration and parallel processing.

Furthermore, LiteMIL’s learnable query vector is not appended to the instance sequence as a [CLS] token, as is common in some vision transformer models. Instead, it operates as a dedicated query that interacts with all instance features. This approach reduces memory usage and computational overhead, as the model does not need to process an augmented sequence or approximate full self-attention over very long input sequences.

TransMIL, a state-of-the-art transformer-based MIL model, employs a [CLS] token and complex attention mechanisms, including Nyström attention and specialized positional encodings, to efficiently handle large bags of instances. While TransMIL achieves strong performance, its architectural complexity and resource demands can be prohibitive in clinical settings with limited computational infrastructure. LiteMIL, by contrast, focuses on architectural simplicity and efficiency, achieving comparable or superior performance with significantly reduced GPU memory usage and training time.

These innovations collectively position LiteMIL as a practical and scalable solution for weakly supervised WSI classification, balancing diagnostic accuracy with resource efficiency and facilitating broader clinical adoption of AI-driven pathology tools.

### 4.2 Impact of Feature Extractor and Bagging Strategy

The integration of Phikon, a pathology-tuned SSL feature extractor, further enhances both performance and efficiency. Compared to conventional CNN-based extractors (e.g., ResNet50), Phikon provides more compact and discriminative representations, leading to an 8% improvement in F1 score and more stable training dynamics. This finding aligns with recent literature emphasizing the superiority of domain-specific SSL models for histopathology tasks (23, 24, 26).

Our chunking strategy for bag formation also contributes to the pipeline’s effectiveness. By partitioning each WSI into multiple uniform bags (in contrast to subsampling, which utilizes only a subset of patches), we increase the number of training samples and preserve local tissue context, which is essential for accurate subtyping. This approach simplifies batch processing, eliminates the need for complex padding or masking, and reduces memory usage (Table 3). This strategy is inspired by pathology practice, where a couple of fields rather than an entire WSI, are sufficient to provide an accurate diagnosis.

### 4.3 Limitations and potential solutions

Despite these strengths, several limitations warrant discussion. First, the TCGA-THYM dataset, while comprehensive, may not fully capture the heterogeneity encountered in routine clinical practice, and class imbalance remains a challenge. Future work should include multi-institutional datasets and explore advanced strategies for handling rare subtypes. Second, while the chunking strategy improves efficiency and data augmentation, it may not optimally capture global context in highly heterogeneous tumors; hierarchical or adaptive bagging approaches could be explored. Third, although the use of SSL features and attention mechanisms improves performance, interpretability remains a concern. Incorporating explainable AI techniques, such as attention heatmaps or saliency maps, could enhance clinical trust and facilitate integration into diagnostic workflows (31).

## 5 CONCLUSION

In summary, LiteMIL offers a practical and scalable solution for cancer subtyping on WSIs, balancing diagnostic precision with computational efficiency. Its streamlined architecture, robust performance across multiple datasets, and reduced resource requirements make it well-suited for deployment in both high- and low-resource clinical settings. Future research should focus on expanding validation to more diverse datasets, integrating multi-modal data, and enhancing model interpretability to further bridge the gap between AI research and clinical practice.

## Statements and Declarations

### Ethics statement

All procedures were performed in compliance with relevant laws and institutional guidelines. The current study is based on public tissue images, and patients’ identities were not disclosed.

### Data Availability

The code to reproduce the study’s results is hosted in a public GitHub repository, accessible at https://github.com/hkussaibi/LiteMIL.

## Acknowledgment

The results shown here are based on data generated by the TCGA Research Network: https://www.cancer.gov/tcga.

## Declaration of generative AI and AI-assisted technologies in the writing process

During the preparation of this work the author(s) used ChatGPT 4.1 in order to improve language and readability. After using this tool/service, the author(s) reviewed and edited the content as needed and take(s) full responsibility for the content of the publication.

